# Top ten research priorities for Essential Emergency Critical Care: A modified Delphi process

**DOI:** 10.1101/2025.09.16.25335940

**Authors:** Ryan Rhys Ellis, Wurry Ayuningtyas, Alexandra Wharton Smith, Martin Gerdin Warnberg, Carl Otto Schell, The EECC Prioritisation Group, Tim Baker

**Author notes:** Corresponding author (TB). Members of the EECC Prioritisation Group are provided in the Acknowledgments.

## Abstract

Critical illness leads to millions of preventable deaths each year. Essential Emergency and Critical Care (EECC) is a pragmatic, globally relevant approach designed to address critical gaps in basic life-saving care. This study identified the top ten research priorities to guide the development of EECC over the next five years. At the first global EECC Research Conference in November 2024, 46 clinicians, researchers, and policy makers participated in a structured priority-setting process. Participants formed four thematic groups focused on the current state, implementation, impact, and refinement of EECC. Throughout five sessions, groups developed sub-themes, generated research questions, and identified key priorities. An anonymous survey was used to rank questions within each theme, followed by a final selection of the top ten. Participants developed 28 research questions across four themes. The final top ten included four on EECC implementation, four on its current state, and two on impact. No top-ranked questions emerged from the refinement theme. The top ten EECC research priorities focused on estimating the burden of critical illness and on implementing programs effectively. Advancing these priorities through collaborative research is essential to strengthen health systems and improve outcomes for critically ill patients globally.

## Introduction

Critical illness has been defined as “a state of ill health with vital organ dysfunction, a high risk of imminent death if care is not provided and the potential for reversibility” and critical care as “identification, monitoring, and treatment of patients with critical illness through the initial and sustained support of vital organ functions”.[1] Patients who are critically ill may be neglected in health systems due to a lack of recognition, resource allocation, prioritisation, and timely identification and treatment with essential life-saving interventions.

Critical illness affects an estimated 45 million adults globally each year, with inadequate access to critical care contributing to millions of preventable deaths.[2] In Africa, for instance, one in eight hospitalised patients is critically ill, and approximately 20% of these patients die within seven days.[3] This is not an isolated issue; similar burdens of critical illness have been documented across diverse geographic and economic contexts, affecting both adults and children.[4] Notably, the majority of critically ill patients are managed in general wards rather than specialised units.[5] In low-resource settings, the availability of even basic supportive care is severely limited, and most critically ill patients do not receive essential life-sustaining care.[6]

Essential Emergency and Critical Care (EECC) has been developed to effectively and pragmatically address the burden of critical illness and move towards ending preventable critical care deaths. EECC is the care that all critically ill patients should receive in all hospitals worldwide, irrespective of their age, gender, diagnosis, or social status. In 2021, 269 global experts from over 50 countries with clinical experience across acute medical specialities took part in a consensus project to reach an agreement on the content of EECC[7], which would be feasible for all hospital wards and units. From this consensus, EECC has started to be integrated into health systems, encouraging a horizontal approach focusing on illness severity rather than speciality and diagnosis to improve care and outcomes for all critically ill patients.[2] Although progress has been made, significant knowledge gaps still exist. As momentum is growing, there is an increasing need for evidence on all aspects of EECC. In this paper, we report the output of an exercise to generate EECC research priorities for the next five years.

## Methods

### Study design

From November 6th to 7th, 2024, the first global research conference on EECC was held at the Department of Global Public Health, Karolinska Institutet, Stockholm, Sweden, in collaboration with Muhimbili University of Health and Allied Sciences, Tanzania. A range of clinicians and researchers were invited to participate, selected based on their research interests and relevant clinical backgrounds. In total, there were 46 participants in the research priority-setting exercise. At the conference, participants were presented with an update on the current state of critical illness, critical care, and EECC research, including insights from previous studies from Africa, Asia, and Europe. A modified Delphi was concurrently held, with participants engaged in structured, interactive group sessions to develop and prioritise EECC research questions.

### Participants

Participants were purposively assigned to one of four groups to achieve an expedient distribution of different areas of expertise. Participants in each group established research priorities in five sequential sessions, each representing one of four designated themes developed by the study leads: i) The current state of EECC; ii) Implementing EECC; iii) The impact of EECC; and iv) Refining EECC. Within each themed group, the members identified the prioritised research questions. The opinions of the gathered experts were gathered via a structured process, described by Schmidt[8], which involved multiple rounds of deliberation leading to a final consensus. This stepwise process progressed through issue generation, preliminary prioritisation, and final ranking via five defined sessions.

### Prioritisation process

The five sessions were conducted over two days with specific objectives to be achieved before progression to the next session. Within each session, a facilitator was assigned to each group to guide the respective thematic areas, and a note-taker was assigned to present the discussions and upload notes and decisions to an online document for analysis. A detailed description of this process is presented below and summarised below (Fig 1).

**Fig 1.**
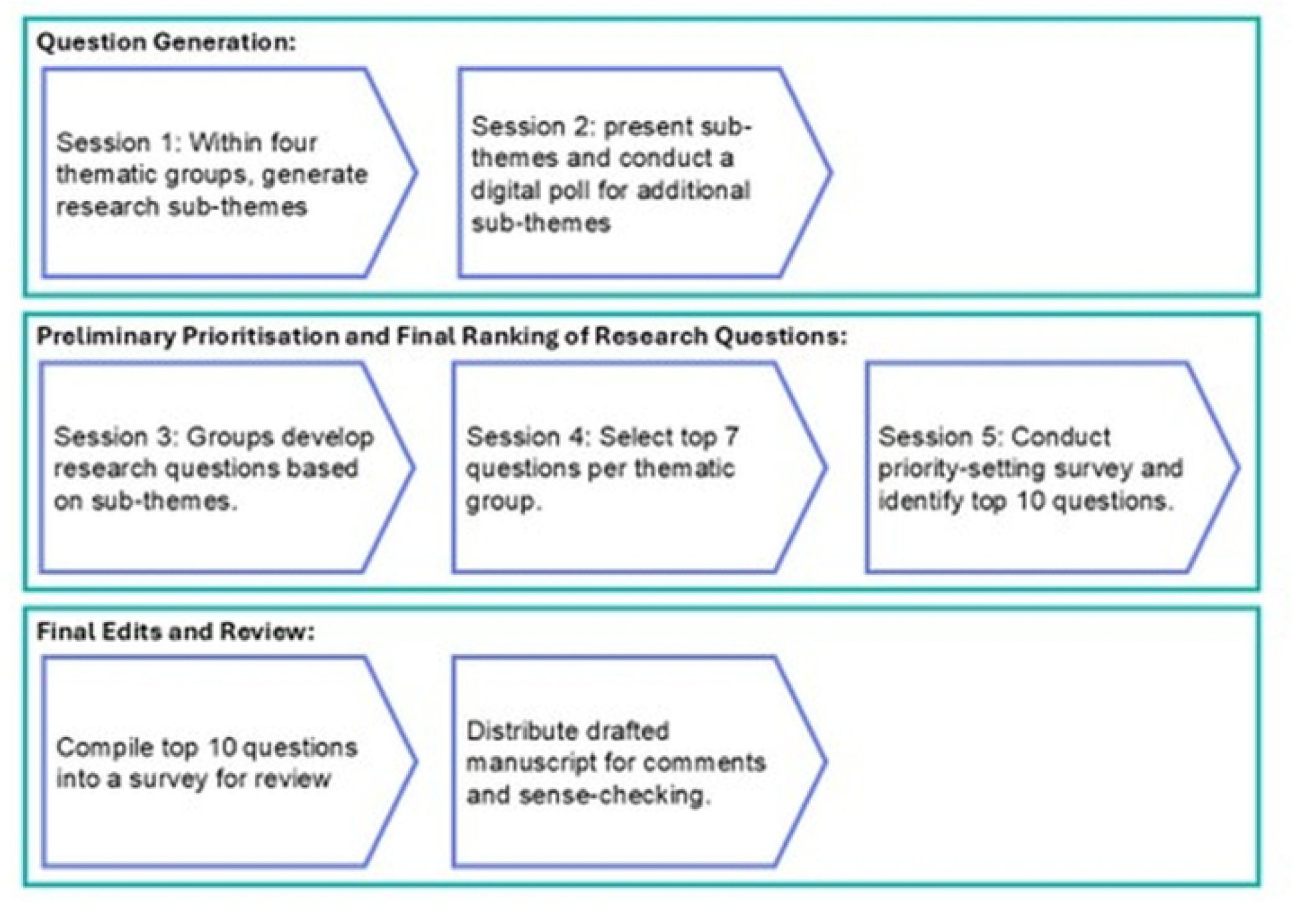
Summary of the priority-setting process

#### Session 1

Each of the four groups was tasked with generating research sub-themes within their designated theme.

#### Session 2

The groups reconvened to present the sub-themes to the entire conference’s participants. An anonymous digital poll was conducted using Menti[9] on participants’ phones or laptops, allowing participants to propose additional relevant sub-themes. The combined input from Sessions 1 and 2 produced an exhaustive list of research sub-themes.

#### Session 3

Participants returned to their groups and developed research questions based on the sub-themes.

#### Session 4

Each group selected the seven most important research questions they had developed, resulting in a total of 28 priority research questions across the four themes.

#### Session 5

All conference participants completed a priority-setting survey using the online platform SmartSurvey.[10] The 28 questions were collectively reviewed, and participants rated each using a 5-point Likert scale ranging from 0 (not important at all) to 4 (essential). The top ten EECC priority questions were identified as those with the highest mean scores across all responses.

### Final Edits and Review

The top ten generated questions were compiled into a survey in SmartSurvey and distributed to the study leads for review, informing the drafting of the manuscript. The drafted manuscript was distributed to all conference participants for further comments and sense-checking.

### Ethics

This study involved only professional participants (clinicians, researchers, and policymakers) contributing their expertise during a structured research priority-setting exercise. No patient data or sensitive personal information was collected. Given the non-clinical and low-risk nature of the activity, formal ethical review was not required. All participants provided informed written consent, and survey responses were collected anonymously.

## Results

Of the 46 participants in the prioritisation process (Table 1), just over half were female (57%, n=26). Physicians constituted the largest occupational group, representing 65% (n=30) of participants, followed by policymakers (7%, n=3), nurses (4%, n=2), and technical advisors (4%, n=2). The majority had research experience, with 20% (n=9) identifying exclusively as researchers. Participants brought a mix of experience from high-income countries (HICs) and low- and middle-income countries (LMICs). At the time of the conference, 50% (n=23) were based in HICs, 17% (n=8) in LMICs, and 33% (n=15) worked across both settings. Nationalities represented included the United States, Nigeria, Tanzania, Ethiopia, Sweden, Norway, Indonesia, the United Kingdom, and New Zealand.

**Table 1.**
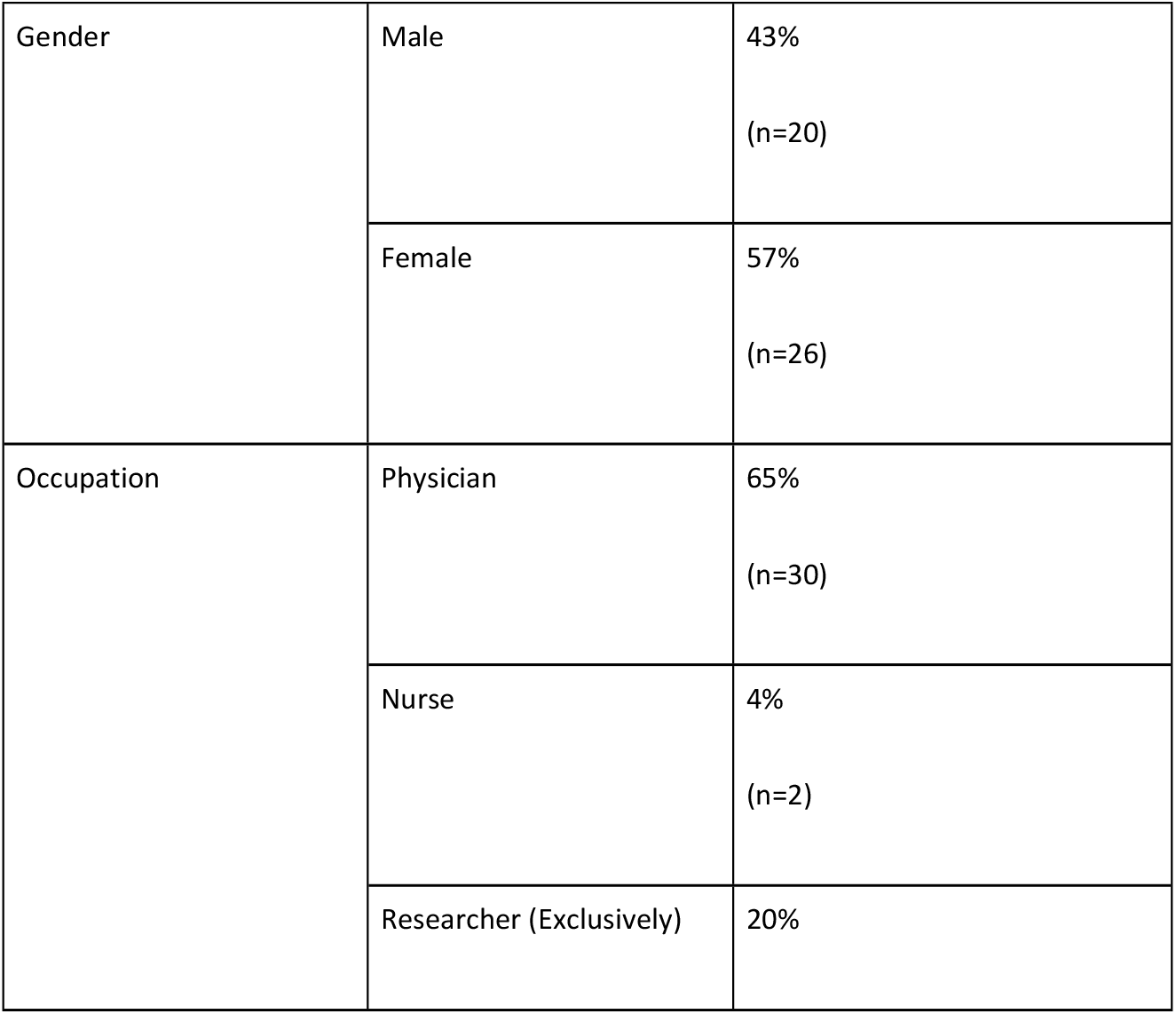
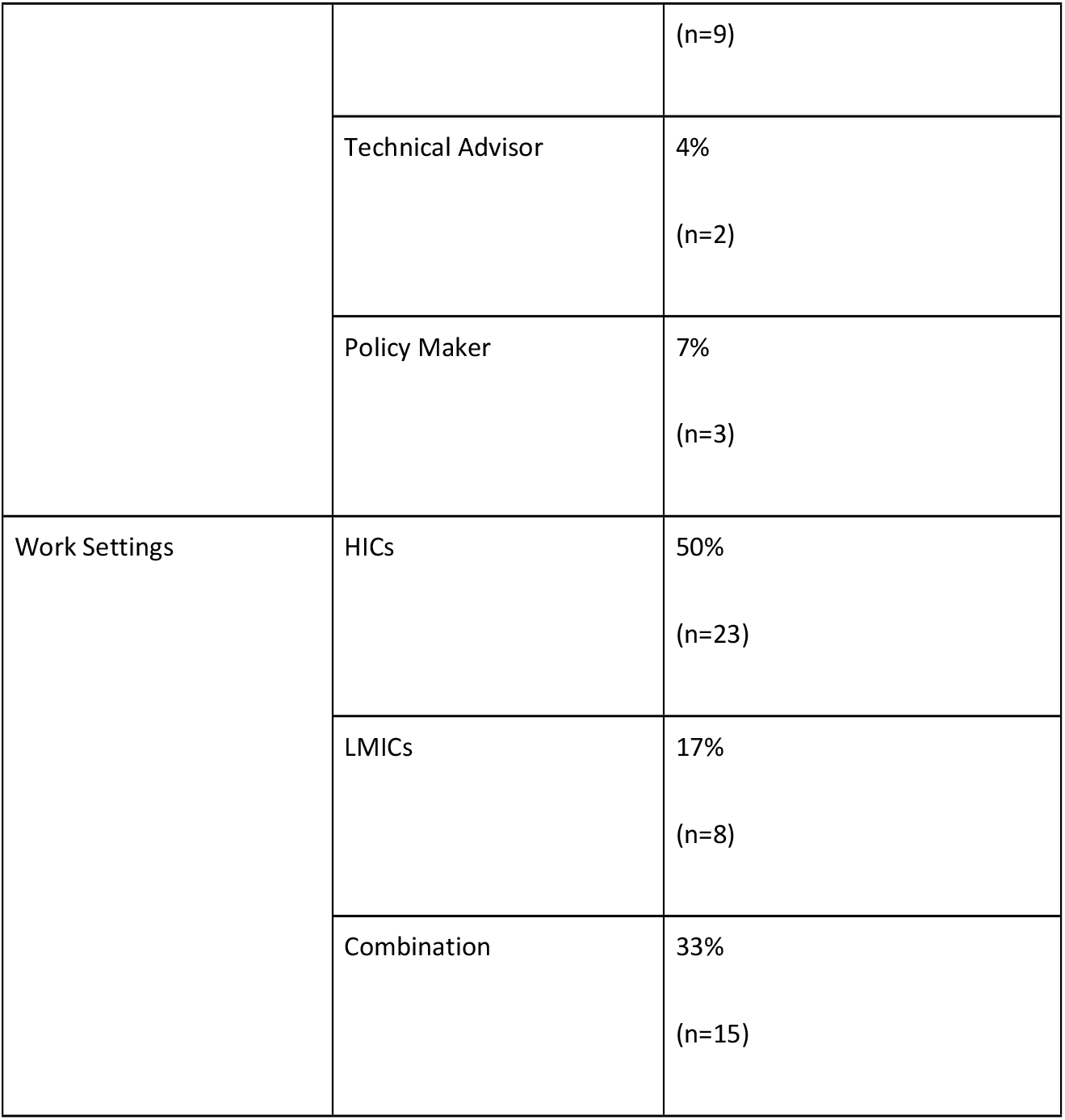
Demographics of conference participants.

The seven priority questions for each of the four themes from Session 4 are presented in Table 2. The final ten prioritised research questions consisted of four questions from the theme of Implementing EECC, four from the Current State of EECC, and two from the Impact of EECC. No questions were chosen from the theme of Refining EECC (Table 3).

**Table 2.** The top seven priority questions from each theme.

|  |  |
| --- | --- |
| **Current State** |  |
| 1. | What is the global burden of critical illness? |
| 2. | What are the best definitions and criteria to measure critical illness? |
| 3. | What are the perceptions about critical illness, critical care, and EECC among different groups (patients, caregivers, health workers, hospital management, health authorities, policy-makers, global stakeholders)? |
| 4. | What are the current provisions for critical care, the existing gaps, prevailing clinical practices, knowledge levels among healthcare providers, and the overall readiness of health facilities to deliver essential critical care services? |
| 5. | What are the ethical considerations, norms and practices around care for critically ill patients? |
| 6. | In selected settings, what is the political economy of EECC (governance, finance, interest groups)? |
| 7. | How might conflict, climate catastrophes, pandemics and public health emergencies impact the critical illness burden and the burden on the health system? |
| **Implementing EECC** |  |
| 1. | What are the underlying determinants of whether EECC is provided or not? |
| 2. | What is the perceived value (cost, appropriateness, acceptability, feasibility) of EECC for stakeholders across different levels? |
| 3. | What are the key indicators of EECC adherence? |
| 4. | What are the facilitators and barriers for healthcare workers to implement and sustain EECC? |
| 5. | How can the components of EECC be operationalised in current policies, guidelines and curricula in different contexts? |
| 6. | What are the core outcomes we should measure in EECC research? |
| 7. | What can we learn from implementing other EECC-related initiatives, implementation models and quality improvement science? |
| **Impact of EECC** |  |
| 1. | What are the impacts of EECC on short and long-term mortality and morbidity? |
| 2. | What is the impact of EECC on outcomes in different patient subgroups? |
| 3. | What is the cost, cost-effectiveness, and budget impact of EECC? |
| 4. | What is the impact of implementing EECC on reducing preventable in-hospital mortality among critically ill patients? |
| 5. | What is the sustainability of EECC implementation? |
| 6. | How does EECC implementation affect patient flows through the health systems? |
| 7 | What are the broader impacts of implementing EECC, including unintentional consequences, for patients, providers, health systems, communities and the environment? |
| **Refining EECC** |  |
| 1. | Should the wording in Essential Emergency and Critical Care be changed to promote greater universal understanding of the concept? |
| 2. | Is a revision of the EECC package necessary at this stage, or should it be scheduled for a future date, and if so, when should this be? |
| 3. | Can AI be used to extract medical records of treatments provided to critically ill patients to inform modifications to EECC? |
| 4. | Which of the 40 defined EECC treatments and actions have the greatest impact on mortality and morbidity, and which are most cost-effective? |
| 5. | Should a reduced EECC package be developed for resource-deficient settings, e.g. pre-hospital and community settings? |
| 6. | Should EECC be differentiated for different population age groups, i.e., neonatal, paediatric and adult? |
| 7. | How best can foundational EECC be linked to more advanced critical care and definitive care programmes? |

**Table 3.**
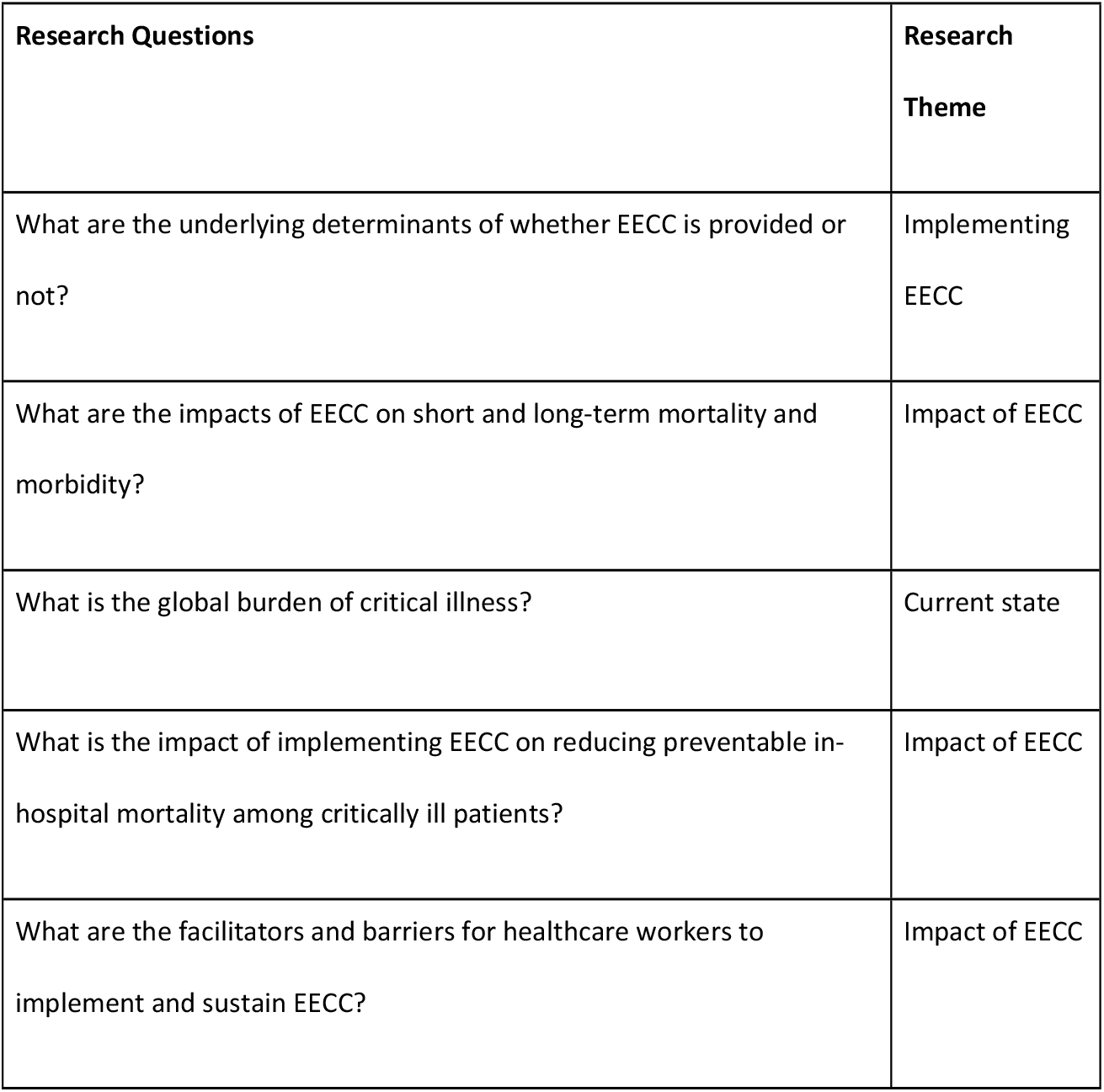
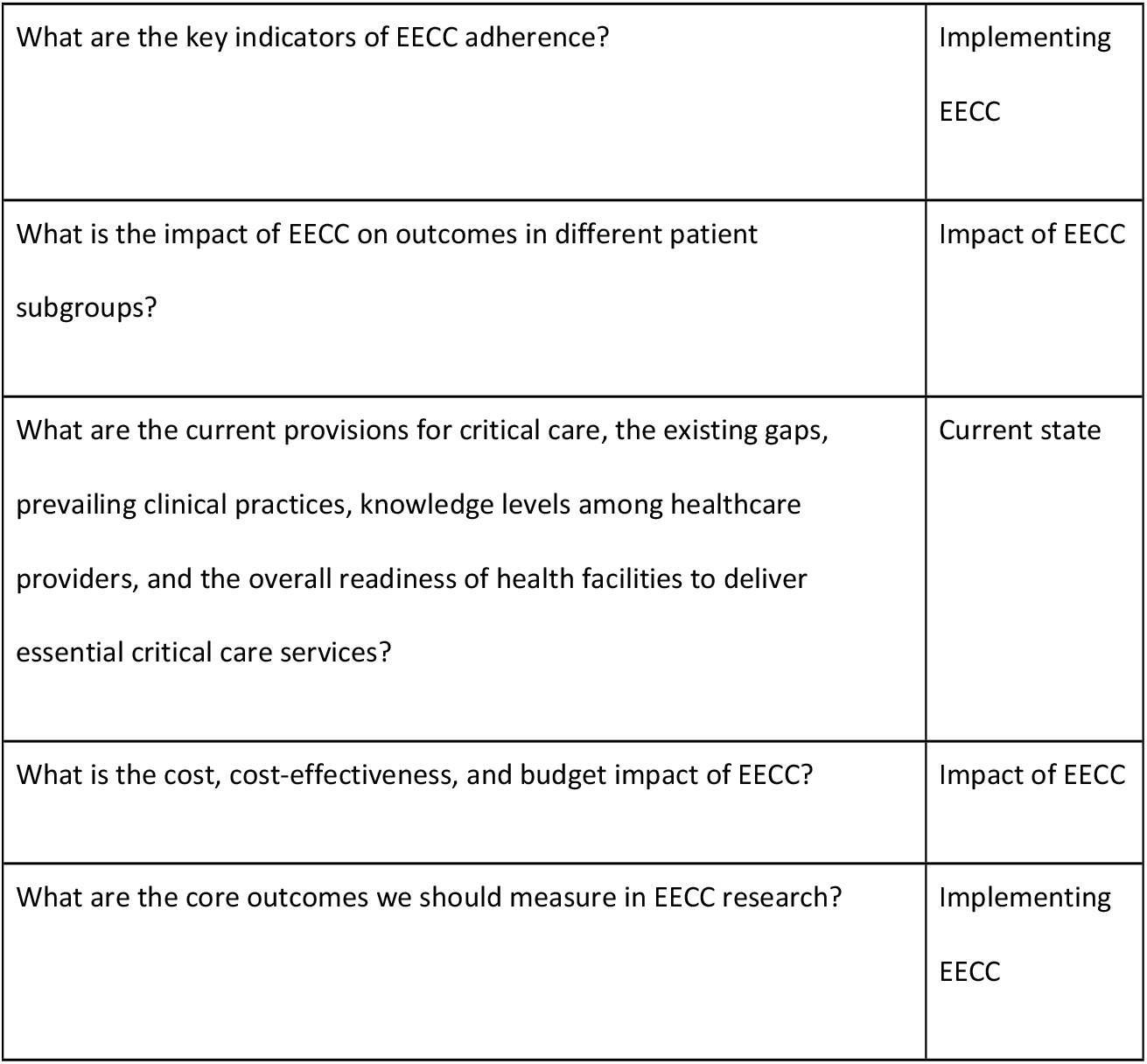
The top ten priority questions.

| **Research Questions** | **Research Theme** |
| --- | --- |
| What are the underlying determinants of whether EECC is provided or not? | Implementing EECC |
| What are the impacts of EECC on short and long-term mortality and morbidity? | Impact of EECC |
| What is the global burden of critical illness? | Current state |
| What is the impact of implementing EECC on reducing preventable in-hospital mortality among critically ill patients? | Impact of EECC |
| What are the facilitators and barriers for healthcare workers to implement and sustain EECC? | Impact of EECC |
| What are the key indicators of EECC adherence? | Implementing<br>EECC |
| What is the impact of EECC on outcomes in different patient subgroups? | Impact of EECC |
| What are the current provisions for critical care, the existing gaps, prevailing clinical practices, knowledge levels among healthcare providers, and the overall readiness of health facilities to deliver essential critical care services? | Current state |
| What is the cost, cost-effectiveness, and budget impact of EECC? | Impact of EECC |
| What are the core outcomes we should measure in EECC research? | Implementing<br>EECC |

## Discussion

Ten prioritised EECC research questions have been developed in this prioritisation process held at the first global EECC research conference. The questions cover areas including the current state of EECC, its implementation, and the impact of EECC provision.

The questions in the current state of EECC focus on the global burden of critical illness and existing provisions and gaps. Recent work has been conducted in Sweden, Sri Lanka, and Malawi[5], as well as across Africa[3]. However, accurate estimates of the global burden of critical illness and the needs are not known. Some geographical areas have been less studied, and EECC research in the Americas, the Middle East, and Asia should be a priority, as well as more granular research in different contexts at the national and local levels.

The implementing EECC questions include the determinants of why EECC is provided or not, the barriers to implementation and sustainability, indicators of adherence, and what core outcomes should be measured when implementing. Sustainable implementation will likely require developments in behavioural change as suggested by the theory of change models[11] that have been successfully used in other settings[12]. Research should investigate how such models and strategies can effectively implement EECC in various contexts and settings. Looking specifically at barriers, previous work on barriers to quality perioperative care delivery in LMIC identified four major themes: fragmented care pathways, limited human and structural resources, the costs of care for patients and patients’ low expectations of care.[13] EECC can address these issues by shifting the focus away from siloed and fragmented care, emphasising the essential resources needed, and advocating for patients’ understanding and right to universal foundational care in critical illness, regardless of the underlying diagnosis.

The successful provision of EECC also depends on the availability of appropriate and context-sensitive training for healthcare providers. An EECC training course[14] has been recently developed, and other important training programs aim to improve emergency and critical care, such as the WHO’s Basic Emergency Care (BEC).[15] Further investigation is warranted to determine how these educational initiatives can be sustainably introduced across varied settings and to assess their impact on provider competencies and patient outcomes. Ultimately, the long-term provision of EECC will require integration with WHO and other stakeholder initiatives, and inclusion in countries’ health systems through a coordinated approach that involves multiple implementation strategies alongside training, including policy development, leadership capacity building, and data collection and evaluation mechanisms to ensure its effectiveness and sustainability across various health system contexts.[16]

Key questions regarding the impact of EECC include its influence on both short- and long-term health outcomes, the proportion of mortality potentially preventable through its implementation, its differential effects across patient subgroups, and its cost-effectiveness and budget implications. Generating robust evidence in these areas is crucial for informing policy decisions and allocating resources effectively. While research into effective implementation strategies is necessary, equal attention must be given to evaluating the impact of EECC. This will require the identification of appropriate outcome measures and the development of rigorous methodological approaches to assess their effectiveness and value across diverse healthcare settings. These goals align with the World Health Assembly Resolution WHA75.8, which establishes a global mandate to strengthen the quality of clinical trials, provide high-quality evidence on health interventions and improve research coordination.[17] The resolution aligns strongly with the need to generate robust, policy-relevant evidence on the effectiveness of EECC implementation, design studies that reflect diverse health systems, and build multidisciplinary networks.

Questions related to the refinement of EECC were not prioritised among conference participants. This may suggest a general acceptance of the current EECC framework, especially given that attendees were already familiar with and had been presented with the existing model. However, this theme may gain prominence in the future, particularly as evolving scientific evidence and technological advancements, including the integration of artificial intelligence, begin to reshape the landscape.

There are several strengths to the methodology used in this research priority-setting process that enhance the validity of the questions generated. The use of a structured, stepwise framework, adapted from the established approach described by Schmidt[8], enabled a systematic progression from question generation to final prioritisation. This design ensured that all participants had multiple opportunities to engage meaningfully throughout the process. The combination of small-group work and plenary sessions promoted collaborative reflection while supporting in-depth, theme-specific exploration. The use of anonymous digital tools (Menti and SmartSurvey) was employed to encourage open and honest input, thereby attempting to mitigate the influence of power dynamics and hierarchical bias, particularly during the ranking phase. A further strength was the inclusion of participants from a range of geographic regions and diverse clinical and research backgrounds, which brought varied contextual perspectives and increased the global applicability of the resulting priorities. Finally, circulating the draft manuscript for participant feedback added a further layer of validation and transparency to the process.

There were also limitations. While the participant group was diverse, encompassing clinical, academic, and policy expertise, it lacked broader representation from other key stakeholder groups, including health system managers, funders, nurses, and patient or community representatives. This limited the scope of perspectives, particularly regarding system-level integration and community impact. Additionally, the geographical scope was relatively narrow. Although participants represented varied economic and global contexts, only nine countries were included, which may limit the diversity of viewpoints. The limited two-day timeframe of the conference further restricted the depth of deliberation, especially for complex or cross-cutting issues. The final ranking process effectively captured the perceived importance of each research question; however, other critical prioritisation criteria, such as feasibility, equity, urgency, and expected impact, were not systematically integrated into the scoring framework. Furthermore, the thematic grouping of participants and questions, while helpful for structuring discussions, may have inadvertently hindered cross-thematic analysis and introduced some redundancy. Finally, although balanced dialogue was supported with facilitators, anonymous polling, and note-takers, the potential influence of dominant voices during early group discussions cannot be entirely discounted.

## Conclusion

Following the first global research conference on Essential Emergency and Critical Care, a set of ten priority research questions has been identified to guide the EECC research agenda over the next five years. These priority questions highlight gaps in knowledge related to the current understanding of burden and care provision, implementation challenges, and the impact of EECC on health outcomes and systems. The next step is to mobilise a diverse group of stakeholders, including researchers, clinicians, policymakers, governments, and funders, to address these priorities collaboratively.

## Data Availability

The manuscript documents the participant demographics and the full list of priorities identified, that was then shortened to a top ten list. No additional data to this was collected.

## Acknowledgements

**The EECC Prioritisation Group members**: Karima Khalid, Dan Brun Petersen, Save Schroder, Kun Arifi Abbas, Septo Sulistio, Alhassan Datti Mohammed, Andreas Barratt-Due, Nobhojit Roy, Andreas Wellhagen, Birger Forsberg, Carina King, Celia Blaas, Christoffer Hintze, Claudia Hanson, Emily Tegnell, Helle Mølsted Alvesson, Jonas Blixt, Louise Elander, Mariam Claeson, Miklos Lipcsey, Petronella Bjurling-Sjöberg, Tobias Alfvén, Aneth Charles Kaliza, Anna Hvarfner, Godfrey Barabona, Ulrika Baker, Nick Leech, Teresa Bleakly Kortz, Halinder Mangat, Jonna idh, and Simon-Fredrik Schell.

## References

1. Kayambankadzanja RK, Schell CO, Wärnberg MG, Tamras T, Mollazadegan H, Baker T, et al. Towards definitions of critical illness and critical care using concept analysis. BMJ Open. 2022 Sep 1;12(9):e060972.

2. Schell CO, Wärnberg MG, Hvarfner A, Höög A, Baker U, Castegren M, et al. The global need for essential emergency and critical care. Crit Care. 2018;22(1):284.

3. Baker T, Scribante J, Elhadi M, Ademuyiwa A, Osinaike B, Owoo C, et al. The African Critical Illness Outcomes Study (ACIOS): a point prevalence study of critical illness in 22 nations in Africa. Lancet. 2025 Mar 1;405(10480):715–24.

4. Kortz TB, Holloway A, Agulnik A, He D, Rivera SG, Abbas Q, et al. Prevalence, aetiology, and hospital outcomes of paediatric acute critical illness in resource-constrained settings (Global PARITY): a multicentre, international, point prevalence and prospective cohort study. Lancet Glob Health. 2025 Feb;13(2):e212–21.

5. Otto SC, Kazidule KR, Abi B, Wärnberg A, Kayambankadzanja C, Hvarfner A, et al. Hospital burden of critical illness across global settings: a point prevalence and cohort study in Malawi, Sri Lanka and Sweden. BMJ Glob Health. 2025;10(3):e017119.

6. Kazibwe J, Shah HA, Kuwawenaruwa A, Schell CO, Khalid K, Tran PB, et al. Resource use, availability and cost in the provision of critical care in Tanzania: a systematic review. BMJ Open. 2022 Nov 1;12(11):e060422.

7. Schell CO, Khalid K, Wharton-Smith A, Oliwa J, Sawe HR, Roy N, et al. Essential emergency and critical care: a consensus among global clinical experts. BMJ Glob Health. 2021 Sep 1;6(9):e006585.

8. Schmidt RC. Managing Delphi surveys using nonparametric statistical techniques. Decis Sci. 1997 Jul;28(3):763–74.

9. Mentimeter AB. Mentimeter [Internet]. Stockholm: Mentimeter AB; [cited 2025 May 15]. Available from: https://www.mentimeter.com/

10. SmartSurvey Ltd. SmartSurvey [Internet]. Tewkesbury (UK): SmartSurvey Ltd.; [cited 2025 May 15]. Available from: https://www.smartsurvey.co.uk/

11. May CR, Mair F, Finch T, MacFarlane A, Dowrick C, Treweek S, et al. Development of a theory of implementation and integration: Normalization Process Theory. Implement Sci. 2009 Dec;4:29.

12. Agulnik A, Ferrara G, Puerto-Torres M, Gillipelli SR, Elish P, Muniz-Talavera H, et al. Assessment of barriers and enablers to implementation of a pediatric early warning system in resource-limited settings. JAMA Netw Open. 2022 Mar 1;5(3):e221547.

13. Bedwell GJ, Dias P, Hahnle L, Anaeli A, Baker T, Beane A, et al. Barriers to quality perioperative care delivery in low- and middle-income countries: a qualitative rapid appraisal study. Anesth Analg. 2022 Dec 1;135(6):1217–32.

14. EECC Global. Training on EECC [Internet]. [cited 2025 Jun 18]. Available from: https://www.eeccglobal.org/training

15. World Health Organization, International Committee of the Red Cross. Basic Emergency Care: Approach to the Acutely Ill and Injured [Internet]. Geneva: World Health Organization; 2018 [cited 2025 Jun 18]. Available from: https://www.who.int/publications/i/item/9789241513081

16. Schell CO, Khalid K, Baker T. Essential emergency and critical care. In: Chopra M, Hou X, Twum Nimako K, Sanam S, editors. Hospitals in Health Systems: Opportunities for Efficient, High-quality, and Integrated Care. Washington (DC): International Bank for Reconstruction and Development / The World Bank; 2024. p.169–77.

17. World Health Organization. Strengthening clinical trials to provide high-quality evidence on health interventions and to improve research quality and coordination. Geneva: World Health Organization; 2022.

